# Donor clonal hematopoiesis and recipient outcomes after transplantation

**DOI:** 10.1101/2021.09.25.21263697

**Authors:** Christopher J. Gibson, Haesook T. Kim, Lin Zhao, H. Moses Murdock, Bryan Hambley, Alana Ogata, Rafael Madero-Marroquin, Shiyu Wang, Lisa Green, Mark Fleharty, Tyler Dougan, Chi-An Cheng, Brendan Blumenstiel, Carrie Cibulskis, Junko Tsuji, Madeleine Duran, Christopher D. Gocke, Joseph H. Antin, Sarah Nikiforow, Amy E. DeZern, Yi-Bin Chen, Vincent T. Ho, Richard J. Jones, Niall J. Lennon, David R. Walt, Jerome Ritz, Robert J. Soiffer, Lukasz P. Gondek, R. Coleman Lindsley

## Abstract

Clonal hematopoiesis (CH) can be transmitted from donor to recipient during allogeneic hematopoietic cell transplantation. Exclusion of candidate donors with CH is controversial since its impact on recipient outcomes and graft alloimmune function is uncertain.

**Methods:** We performed targeted error-corrected sequencing on samples from 1727 donors aged 40 or older and assessed the effect of donor CH on recipient clinical outcomes. We measured long-term engraftment of 102 donor clones and cytokine levels in 256 recipients at 3 and 12 months after transplant.

**Results:** CH was present in 22.5% of donors, with *DNMT3A* (14.6%) and *TET2* (5.2%) mutations being most common; 85% of donor clones showed engraftment in recipients after transplantation, including clones with variant allele fraction (VAF)<0.01. *DNMT3A-*CH with VAF≥0.01, but not smaller clones, was associated with improved recipient overall (HR 0.79, P=0.042) and progression-free survival (HR 0.72, P=0.003) after adjustment for significant clinical variables. In patients receiving calcineurin-based GVHD prophylaxis, donor *DNMT3A-*CH was associated with reduced relapse (sHR 0.59, P=0.014), increased chronic GVHD (sHR 1.36, P=0.042), and higher IL-12p70 levels in recipients. No recipient of sole DNMT3A or *TET2*-CH developed donor cell leukemia (DCL). In 7 of 8 cases, DCL evolved from donor CH with rare *TP53* or splicing factor mutations or from donors carrying germline *DDX41* mutations.

**Conclusion:** Donor CH is associated with clinical outcomes in transplant recipients, with differential impact on alloimmune function and potential for leukemic transformation related to mutated gene and clonal abundance. *DNMT3A*-CH is associated with improved recipient survival due to reduced relapse risk and an augmented network of inflammatory cytokines in recipients. Risk of DCL is driven by pre-existing somatic MDS-associated mutations or germline predisposition in donors.

## Introduction

Allogeneic hematopoietic cell transplantation is the only curative treatment for most high-risk hematologic malignancies. Patients with an available donor have improved overall survival compared with patients who do not have a donor.^1,2^ The National Marrow Donor Program (NMDP) prioritizes donors under age 45^3^ because younger donor age has been associated with improved recipient survival,^4^ but suitable younger donors are not uniformly available to all transplant candidates. The likelihood of finding a matching donor in the NMDP Be The Match Registry varies widely depending on ethnic background. While HLA-matched adult donors can be identified in the NMDP registry for 75% of Caucasian patients, far fewer patients of color will find matched donors (16-19% of African Americans; 33-42% of Asian Americans, and 34-40% of Hispanic Americans).^5^ The best available graft for many patients may thus be from an older donor such as an HLA-identical sibling or haploidentical relative,^6,7^ highlighting the importance of understanding characteristics of older donors that influence recipient outcomes.

Clonal hematopoiesis is an age-related, asymptomatic condition in which leukemia-associated somatic mutations are detected in the blood of individuals without a hematologic malignancy. In the non-transplant setting, CH is uniformly associated with adverse outcomes, including an elevated risk of developing hematologic malignancies^8,9^ and an increased risk of non-hematologic outcomes due to altered inflammatory signaling.^10^ These clinical effects of native CH are apparent with large clones,^8,10^ but CH at very low clonal abundance with uncertain biological effect has been reported to be ubiquitous in adults over age 40^11^

Exploratory studies have found that CH in transplant donors can engraft in recipients^12–15^ but reported conflicting results on the impact of donor CH on transplant-specific clinical outcomes, graft immunologic function, or risk of donor cell leukemia.^12,13,16,17^ These studies have been limited by modest sample sizes that impacted outcomes analysis, cohort characteristics that restricted generalizability, and a lack of mechanistic rationale. Furthermore, these cohorts have not parsed the clinical impact of different CH mutations or used sequencing technologies that support evaluation of low-abundance clones, which could have unique dynamics in the context of allogeneic transplantation. Current evidence has thus been insufficient to resolve disagreement about whether to screen older candidate donors for CH,^18,19^ and some transplant centers have begun excluding donors found to have CH based on the assumption that the adverse associations of native CH also apply in the context of transplant.^20^ In this study, we performed a comprehensive analysis of samples from donors age 40 or older to determine the impact of clonal hematopoiesis on overall recipient outcomes, risk of donor cell leukemia, and measures of graft alloimmune activity.

## Results

### Clinical and genetic characteristics of donor CH

We identified CH with VAF ≥0.005 in 388 of 1727 donors (22.5%). Its prevalence increased with advancing age: CH was present in 12.6% of donors aged 40-49, 26.6% of donors aged 50-59, and 41.2% of donors aged 60 or older. The proportion of donors with CH in each age decade was similar in both cohorts (Figure 1A). In multivariable analysis stratified by cohort and considering donor, recipient, and transplant variables, only older donor age was independently associated with the presence of CH (median 56 versus 49 years with and without CH, respectively, P<0.001, Table 1).

**Figure 1.**
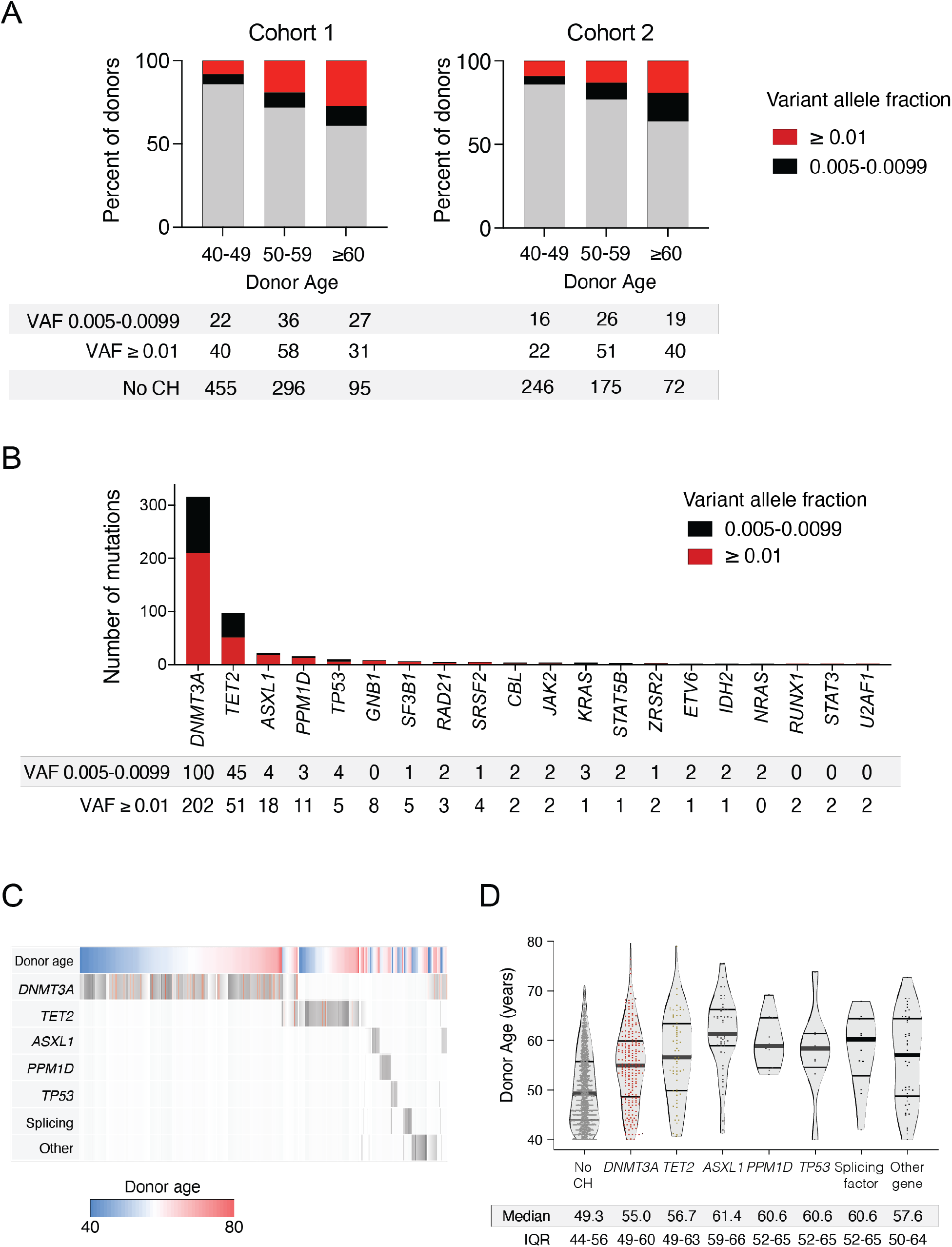
Characteristics of CH in transplant donors aged 40 and older. Panel A shows the proportion of donors with and without CH in each cohort, subdivided by donor age decade. CH with variant allele fraction (VAF) 0.005-0.0099 is in black, and CH with VAF ≥0.01 is in red. Panel B shows the number of mutations in each gene mutated in 2 or more donors, with variants at VAF 0.005-0.0099 again in black and variants at VAF ≥0.01 in red. Panel C shows the patterns of co-mutation among donors with CH. Each column represents a donor, with rows for donor age and the most commonly mutated genes or gene groups. Donors were hierarchically grouped based on the presence of mutations in genes other than *DNMT3A* or *TET2*, then *DNMT3A*, then *TET2*. Genes mutated more than once in the same donor are in red. Panel D shows the distribution of donor ages based on CH status and mutations in individual genes or groups of genes. Medians and interquartile ranges (IQRs) are reported below each column.

**Table 1.**
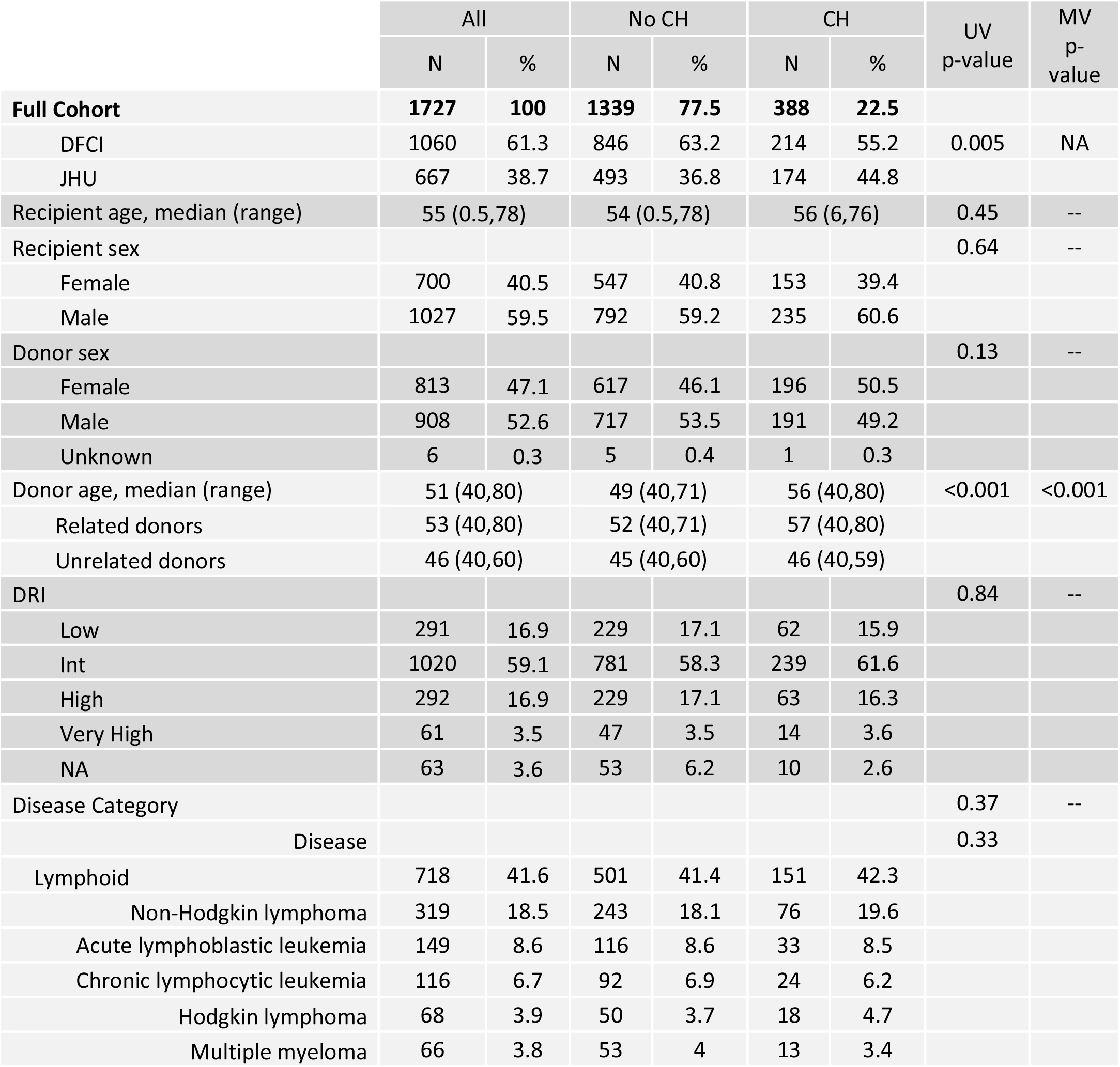
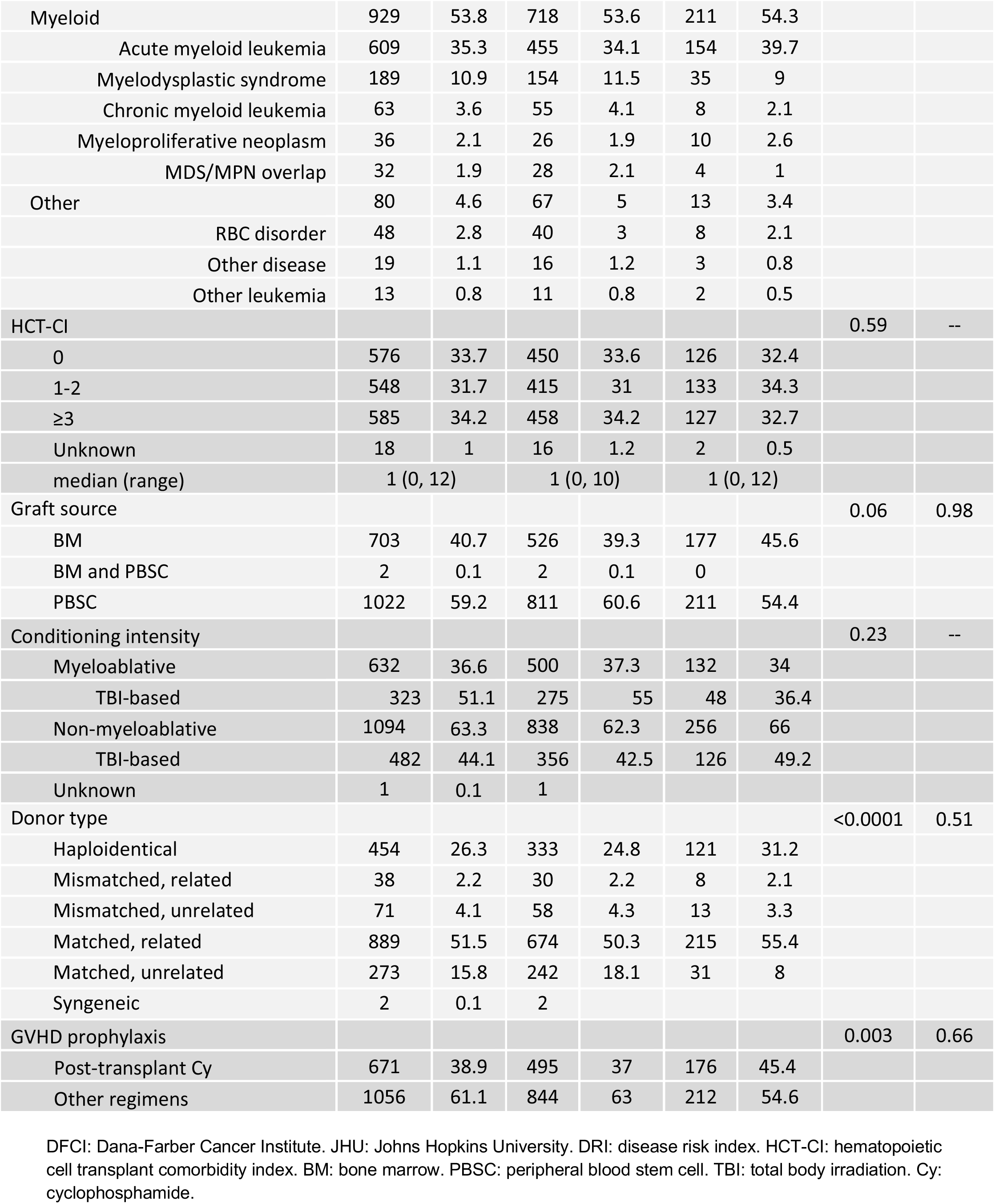
Characteristics of donors and recipients of donors with and without clonal hematopoiesis (CH). The top row reports the proportion of donors with and without CH and sums to 100. Otherwise, the distribution within each category sums to 100 within columns. Univariable (UV) comparisons between those with and without CH were performed with Chi-square or Fisher’s exact test for categorical variables and the Wilcoxon rank-sum test for continuous variables. All variables with UV P-values less than 0.2 were tested for association with presence of CH in a multivariable (MV) model stratified by center.

Among 501 total mutations, 324 had VAF ≥0.01 and 177 had VAF 0.005-0.0099. The most frequently mutated genes were *DNMT3A* (302 mutations in 253 donors), *TET2* (96 mutations in 89 donors), *ASXL1* (22 mutations in 22 donors) and *PPM1D* (14 mutations in 14 donors, Figure 1B). No other gene was mutated in more than 10 donors. Most donors with CH (n=301, 77.5%) had only one mutation. Donors with non-*DNMT3A/TET2* mutations were more likely to have mutations in more than one gene (compared with donors who had *DNMT3A* or *TET2* mutations, OR 3.5, 95% CI 2.0-6.1, P<0.0001, Figure 1C). Donors with CH were older than those without CH irrespective of the gene mutated (Figure 1D). Using an orthogonal duplex UMI-based sequencing platform we validated 100% (28 of 28) variants from 20 donors (VAF range 0.0056 - 0.2159)(Figure S1).

### Donor CH and recipient outcomes

In native CH, larger clones have a greater effect on clinical outcomes than smaller clones,^8,10^ but no evidence-based VAF cutoff for defining CH has been established. To define a clinically relevant VAF threshold for CH in the setting of transplantation, we examined the relative hazards of PFS, relapse, and non-relapse mortality across the full range of donor clone size and found that the effect of CH was greatest with VAF ≥0.01 (Figure S2). Recipients of donor CH with VAF ≥0.01 (n=241) had improved PFS compared with recipients whose donors did not have CH in a multivariable model that included recipient and donor age, donor/recipient sex mismatch, hematopoietic cell transplantation-specific comorbidity index score,^21^ conditioning intensity, donor type, disease category, and Disease Risk Index classification^22^ (HR 0.79, 95% confidence interval [CI] 0.66-0.95, P=0.011, Figure 2A). CH with only smaller donor clones (VAF 0.005-0.0099, n=147) was not significantly associated with any outcome.

**Figure 2.**
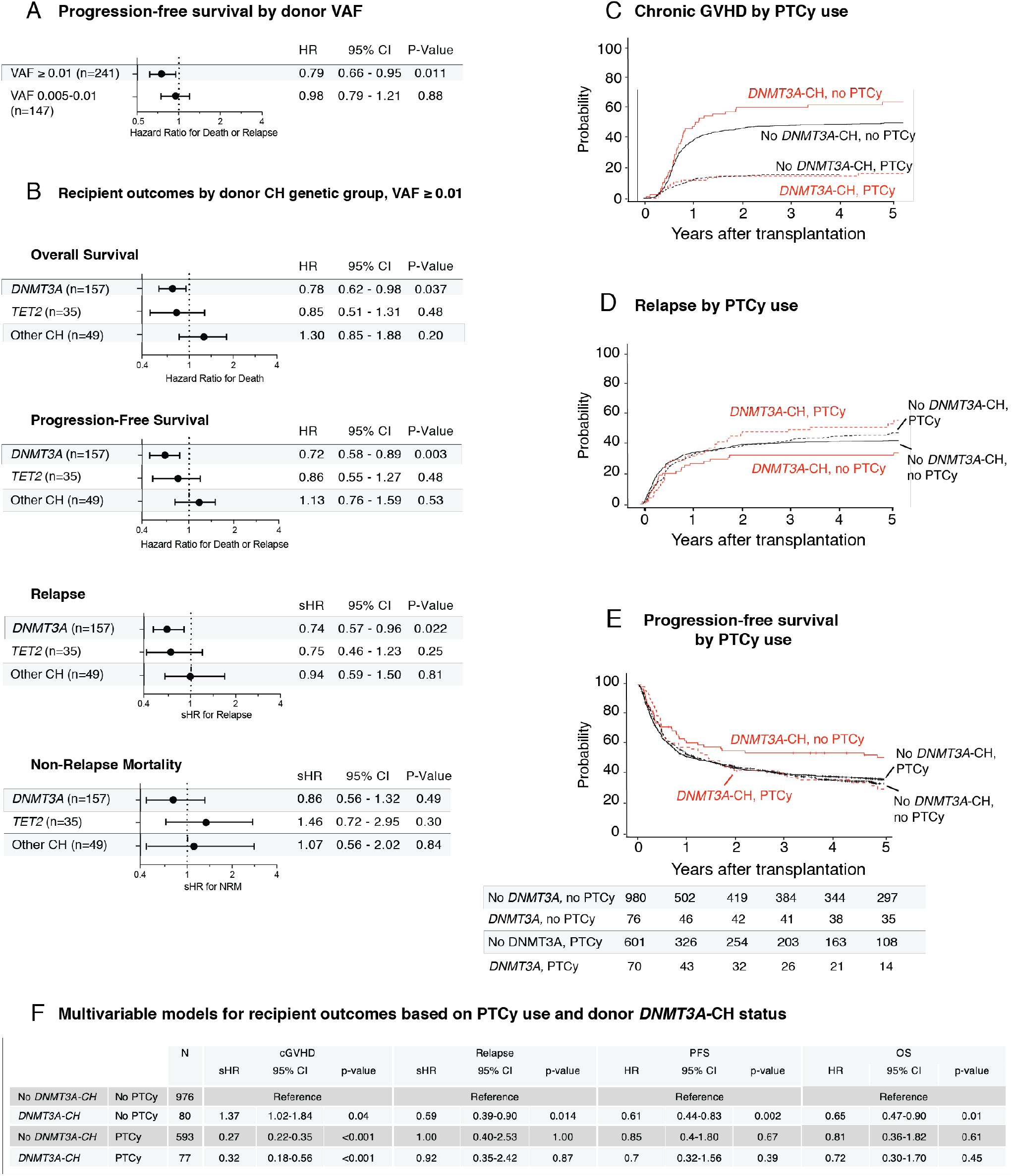
Association of donor CH with recipient clinical outcomes. Panel A shows the association between donor CH at either VAF 0.005-0.0099 or VAF ≥0.01 and recipient progression-free survival (PFS) in multivariable Cox proportional hazard models. Panel B shows the association between donor CH at VAF ≥0.01 and recipient overall survival, PFS, relapse, and non-relapse mortality, in multivariable models divided by gene group. The groups consist of CH with any mutation other than *DNMT3A/TET2* (“Other CH”), then remaining *DNMT3A* mutations, then remaining *TET2* mutations. Panel C shows the 5-year cumulative incidence of chronic graft-versus-host disease (GVHD) split by *DNMT3A-*CH status (VAF ≥0.01) and receipt of post-transplant cyclophosphamide (PTCy) for GVHD prophylaxis. Panel D shows the 5-year cumulative incidence of relapse split by *DNMT3A-*CH status (VAF ≥0.01) and receipt of PTCy. Panel E shows 5-year PFS split by *DNMT3A-*CH status (VAF ≥0.01) and receipt of PTCy. Panel F summarizes the associations between *DNMT3A-*CH and outcomes among recipients who did or did not receive PTCy in multivariable Fine-Gray competing risk regressions (chronic GVHD, relapse) and Cox proportional hazards models (PFS and OS).

Individual gene mutations in CH have distinct associations with clinical outcomes. In non-transplant settings, *DNMT3A* and *TET2* mutations have driven the bulk of association with inflammatory outcomes like cardiovascular disease,^10^ while non-*DNMT3A/TET2* mutations have been associated with a higher risk of progression to hematologic malignancy.^10,23,24^ We therefore tested the effects of three pre-specified, hierarchically defined groups at VAF ≥0.01: (1) donors with mutations in any gene other than *DNMT3A* or *TET2*, (“Other CH,” n=49); (2) remaining donors with *DNMT3A* mutations (“*DNMT3A-*CH,” n=157); (3) remaining donors with only *TET2* mutations (“*TET2-*CH,” n=35).

Only donor *DNMT3A-*CH was significantly associated with recipient outcomes. Recipients whose donors had *DNMT3A*-CH had improved OS and PFS and reduced risk of relapse in the same multivariable model (HR for death 0.78, 95% CI 0.62-0.98, P=0.037; HR for death or relapse 0.72, 95% CI 0.58-0.89, P=0.003; sHR for relapse 0.74, 95% CI 0.57-0.96, P=0.022, Figure 2B and table S6), when compared with recipients whose donors did not have CH with VAF ≥ 0.01. *DNMT3A-*CH was not associated with differences in non-relapse mortality, and causes of death without relapse were similar irrespective of the presence of *DNMT3A*-CH (Table S7). Neither *TET2-*CH nor Other CH had significant impacts on outcomes.

Alloreactive donor immune cells reduce relapse via graft-versus-leukemia activity, but also mediate a complementary risk of GVHD. Conventional GVHD prophylaxis with calcineurin-based regimens suppresses global T cell function, while post-transplant cyclophosphamide (PTCy) is thought to prevent GVHD by selective suppression or elimination of pathogenic alloreactive T cells.^25,26^ To evaluate the interactions between *DNMT3A*-CH and immune-modulating therapy, we analyzed GVHD outcomes in patients who did or did not receive PTCy for GVHD prophylaxis. In multivariable analysis, *DNMT3A-*CH with VAF ≥0.01 was independently associated with an increased risk of chronic GVHD in recipients who did not receive PTCy (Figure 2C; sHR 1.37, 95% CI 1.02-1.84, P=0.04 in multivariable analysis). In contrast, we observed no effect of *DNMT3A*-CH on chronic GVHD in recipients who received PTCy (compared with PTCy and no *DNMT3A-*CH, sHR 1.15, 95% CI 0.82-1.6, P=0.88). The improvements in relapse, PFS, and OS associated with *DNMT3A*-CH were also confined to recipients who did not receive PTCy (Figure 2D-F and table S8). Among these patients, donor *DNMT3A-*CH was associated with reduced risk of relapse (sHR from multivariable model 0.59, 95% CI 0.39-0.9, P=0.014) and reduced risk of death (HR 0.65, 95% CI 0.47-0.9, P=0.01). This difference was evident for both myeloid and lymphoid diseases (Figure S3), and was not evidently influenced by either bone marrow graft source (Figure S4) or haploidentical donors (Tables S9-10), both of which are closely associated with PTCy use. There was no association between *DNMT3A*-CH and acute GVHD with or without PTCy (Figure S5).

### Engraftment and biologic characteristics of donor CH in recipients

We next sought to define the capacity of donor clones to contribute to long-term hematopoiesis in recipients. We sequenced samples collected 3 and 12 months after transplant from 69 recipients whose donors had CH and who survived without relapse for at least one year (Table S11). In total, 86 of 102 donor mutations were detectable in paired recipients at 12 months (Figure 3A), including 84.9% of *DNMT3A* mutations, 94.4% of *TET2* mutations, and 70.6% of other mutations (Figure 3B). Mutations with VAF ≥0.01 engrafted more frequently than mutations with VAF <0.01 (91.7% vs. 76.2%, P=0.045, Figure S6). No other donor, recipient, or transplant variables were associated with the likelihood of 12-month engraftment of the clone (Table S12).

**Figure 3.**
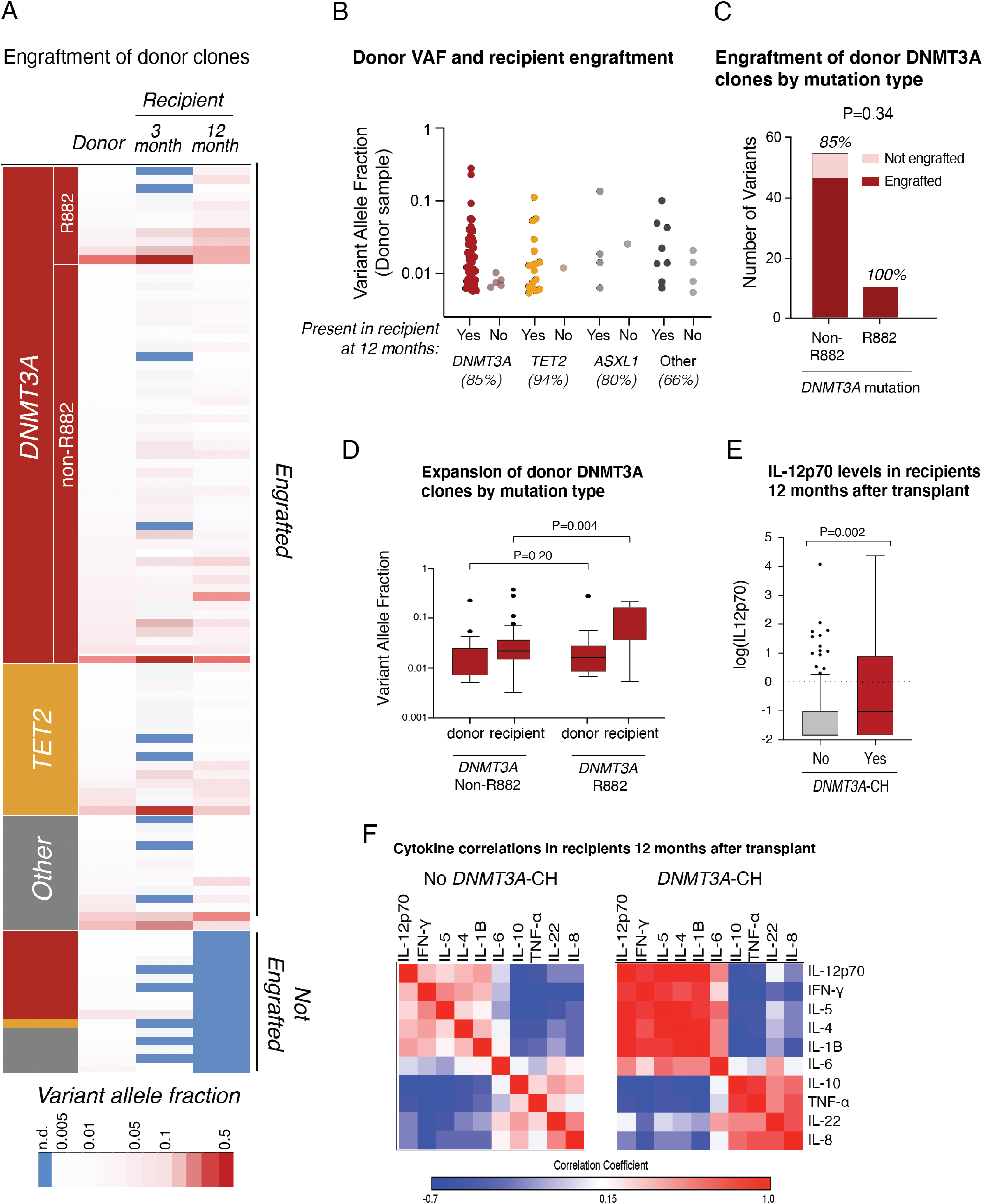
Engraftment of donor CH in recipients after transplantation. Panel A shows the VAFs of 102 donor mutations assessed in 69 recipients following transplantation. Each row is an individual mutation, with columns representing the VAFs in the donors at the time of transplant and in the recipients at 3 and 12 months after transplant. VAFs are color-coded from white (lower VAF) to red (higher VAF), with blue indicating that the mutation was not detected. Mutations that were detectable in recipients at 12 months (engrafted) are on top and those that were not engrafted are on bottom. Panel B shows the donor VAFs of *DNMT3A, TET2, ASXL1*, and other mutations that did or did not engraft in recipients. Panel C shows the proportion of R882 and non-R882 *DNMT3A* mutations assessed post-transplant (n=10 and 54, respectively) that did or did not engraft in recipients at 12 months. Panel D shows the VAFs of non-R882 (left) and R882 *DNMT3A* mutations (right) in donors and corresponding 12-month samples from recipients. Panel E shows the plasma levels of IL-12p70 at 12 months after transplant in recipients of *DNMT3A-*CH (n=21, red) compared with other recipients (n=241, gray). Panel F shows correlations between 10 plasma cytokines measured at 12 months after transplant in the same recipients with and without *DNMT3A-*CH. Correlations are color-coded from blue (negative correlation) to red (positive correlation).

*DNMT3A* encodes the methyltransferase responsible for *de novo* DNA methylation in hematopoietic stem cells,^27^ and a broad spectrum of mutations that variably reduce DNMT3A enzymatic function are frequently identified as early events in myeloid malignancies.^28–30^ Since *DNMT3A* R882 hotspot mutations have distinct biochemical function and higher risk of progression to acute myeloid leukemia compared with non-R882 *DNMT3A* mutations,^31–34^ we further analyzed the long-term engraftment and expansion of 10 R882 and 54 non-R882 clones (median baseline VAF 0.017 versus 0.012, P=0.20). All 10 donor R882 mutations were detectable in recipients at 12 months, compared with 46 of 54 non-R882 mutations (85.2%, P=0.34, Figure 3C). At 12 months, the VAF of R882 mutations was significantly higher than non-R882 mutations (median 0.051 versus 0.021, P=0.004, Figure 3D, Table S13).

In native clonal hematopoiesis, *DNMT3A* mutated stem cell clones invariably contribute to myeloid differentiation, but have also been reported to contribute to mature lymphoid cells.^11,35^ The lineage potential of allogeneic donor-engrafted clones, however, has not previously been assessed. To determine whether *DNMT3A* clones from older donors contribute stably to the T cell lineage after transplantation, we quantified the representation of 19 *DNMT3A* mutations from 14 donors in purified recipient CD3-positive blood cells one year after transplantation. We found that 18 of 19 (94.7%) donor *DNMT3A* mutations were detectable, including 14 of 15 (93.3%) of clones from patients treated with PTCy (table S14).

Native CH has been associated with alterations of inflammatory cytokines,^10^ which could modulate graft immunologic function^36,37^ or accelerate age-associated phenotypes of hematopoietic stem cells from older donors by altering the bone marrow microenvironment.^38^ We assessed the association of donor CH with levels of 10 cytokines in 256 recipients with (n=54) or without (n=202) donor CH who were alive without relapse at 12 months after transplant. Recipients of donor *DNMT3A-*CH at VAF ≥ 0.01 (n=21) had higher median IL-12p70 levels (0.37 pg/ml) than other recipients (n=241, 0.16 pg/ml, Figure 3E, P=0.002). In *DNMT3A-*CH recipients, IL-12p70 levels were positively correlated with levels of IL-1B, IL-4, IL-5, and IFN-*γ*, and negatively correlated with IL-8, IL-22, TNF-α, and IL-10 (Figure 3F, Tables S15-16).

### Evolution to Donor Cell Leukemia

The evolution of donor CH to donor cell leukemia has been described in individual cases,^39–41^ but the leukemic risk of donor CH has not been systematically evaluated in a single cohort. We identified 8 cases of donor cell leukemia, for a 10-year cumulative incidence of 0.7% (Figure 4A). The median latency between transplantation and donor cell leukemia diagnosis was 5.2 years (range 0.3-10.3 years). Donors for recipients who developed donor cell leukemia were significantly older than donors for recipients who did not develop donor cell leukemia (median age 57.3 versus 50.9 years, P=0.037, Figure 4B).

**Figure 4.**
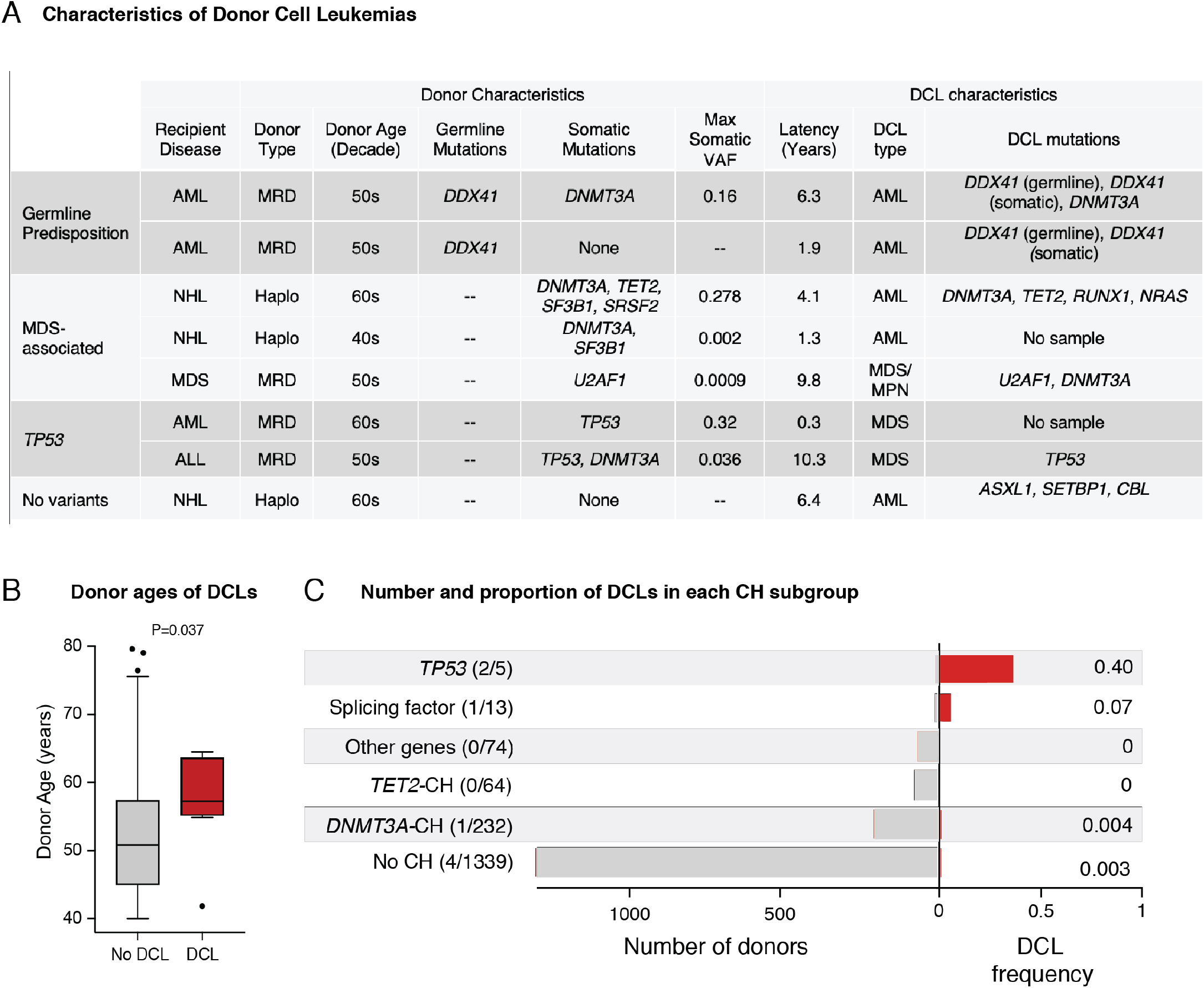
Clinical and genomic features of donor cell leukemias. Panel A shows the clinical and genomic features of 8 donor cell leukemias that developed in the cohort during the study period. Recipient diseases: acute myeloid leukemia (AML), non-Hodgkin lymphoma (NHL), myelodysplastic syndrome (MDS), acute lymphoblastic leukemia (ALL). Donor cell leukemia type: AML, MDS, or MDS/myeloproliferative neoplasm overlap syndrome (MDS/MPN). Somatic mutations reported include both those meeting criteria for CH and lower-abundance mutations identified by sequencing the donor cell leukemia sample. For donor samples with multiple somatic mutations, the maximum VAF is reported. Mutations present in the donor cell leukemia are reported for the 6 of 8 donor cell leukemias with available samples. Panel B shows the distribution of donor ages for recipients who did or did not develop donor cell leukemia. Panel C shows the total number of DCLs in each genetic subgroup of CH on the left, and the corresponding proportion of recipients in each group who developed donor cell leukemia on the right. Note that CH is defined here as VAF ≥ 0.005, meaning two of the donors with low-abundance splicing factor mutations reported in 4A are classified as “no CH.”

To determine the proportion of donor cell leukemias that evolved directly from donor CH, we sequenced six donor cell leukemia samples and analyzed the correlation between donor cell leukemia mutations and the mutations present in the donor. In 5 of 6 cases, donor cell leukemia mutations were detectable in the donor sample. In both donor cell leukemias without available samples, the donors had detectable CH. Donor clones that progressed to leukemia were genetically distinct from more common *DNMT3A* or *TET2* mutated CH: 3 had mutations in MDS-associated splicing factors (*SF3B1/SRSF2/U2AF1*),^42^ 2 had *TP53* mutations, and in 2 cases we identified germline mutations in the leukemia predisposition gene *DDX41* that were shared between recipient and their related donors (Figures S7-S9).^43^ Although donor CH with splicing factor or *TP53* mutations were infrequent in the overall cohort, relatively large proportions developed donor cell leukemia (Figure 4C).

## Discussion

Clonal hematopoiesis in older stem cell donors may influence recipient outcomes after allogeneic transplantation,^13^ but previous studies have not shown an association between donor CH and overall recipient survival, examined its genetic heterogeneity and interaction with immune-modulating therapies, or evaluated the impact of low-abundance clones. Here, we paired sensitive detection of clonal hematopoiesis in 1727 healthy older transplant donors with comprehensive clinical annotation of recipient outcomes. Our findings directly inform screening and selection of older donors and have implications for our fundamental understanding of the graft-versus-leukemia (GVL) effect. We find that the presence of *DNMT3A-CH* or *TET2-CH* CH in stem cell donors does not adversely affect recipient outcomes, and that *DNMT3A-*CH is independently associated with improved survival in recipients as a consequence of reduced relapse. In contrast, rare mutations in MDS-associated genes pose a high risk of leukemic transformation following transplant.

Concerns about the risks of donor clonal hematopoiesis have stemmed largely from extrapolating results of non-transplant studies linking CH to increased risks of hematologic malignancies^8^ and non-hematologic diseases.^10^ These effects are reported to be caused by pathologic dysregulation of inflammatory cytokine signaling from terminally differentiated clonal myeloid cells. In this study, we found that recipients of donor CH had similar evidence of altered inflammatory cytokine signaling, but paradoxically had improved survival mediated by a lower risk of disease relapse, consistent with the critical importance of graft-versus-tumor immunity in allogeneic transplant efficacy. Increased inflammatory signaling from *DNMT3A-*mutant myeloid cells could augment graft alloimmune activity by direct effects on T cell function,^44^ or by effects on malignant cells, where inflammatory signaling positively regulates MHC Class II expression that is required for maintaining tumor immunogenicity after transplantation.^45^ IL-12p70, which was significantly higher in recipients of *DNMT3A-*CH than others and was correlated with other pro-inflammatory cytokines, has been specifically implicated in the development of GVHD and GVL through its positive effects on Th1 polarization and IFN-g production in CD4+ T-cells.^44,46–48^ Crosstalk with donor-engrafted myeloid cells might explain why *DNMT3A-*CH is associated with an increased risk of chronic GVHD, but not acute GVHD, which is mediated by alloreactive T cells present in the graft at the time of transplantation.^49^ Loss-of-function *DNMT3A* mutations in donor-engrafted T cells could also potentiate alloimmune activity in a cell autonomous fashion by limiting exhaustion programs^50^ or augmenting development of memory CD8+ T cell populations,^51^ both of which are mediated by *de novo* DNA methylation in T cells. Together, our results provide a mechanistic rationale for exploring therapeutic modulation of DNA methyltransferase activity to augment efficacy of cell-based immune therapies, thereby improving the antileukemic effect and cure rate in patients with hematologic malignancies.

We empirically defined a VAF cutoff for CH clinical significance. We found that even the smallest clones engraft reliably in recipients, but that the biological and clinical consequences of donor *DNMT3A*-CH are only meaningful above VAF 0.01. Notably, 246 of 388 donors with CH had VAF 0.02 and thus would not have been evaluated in studies relying on the provisional definition of clonal hematopoiesis of indeterminate potential (CHIP).^13^ This is particularly important for clinical applications, as small clones are expected to be identified more frequently as ever more sensitive sequencing technologies are deployed in the clinical setting.

The potential for leukemic evolution of pre-existing donor clones has been proposed as a basis for excluding candidate donors with CH,^18,20,39^ but the magnitude of risk based on CH genotype has not been defined. In our study, no recipients of *DNMT3A* or *TET2* mutated donor CH developed donor cell leukemia without concurrent MDS-associated mutations or germline risk alleles. Instead, donor cell leukemias arose from donor gene mutations that were rare in the cohort overall, such as *TP53* and MDS-associated splicing factors, supporting findings of published case reports.^39,52,53^ High-risk mutations were also associated with older donor age, raising the possibility that age alone also contributes to risk of transformation. The frequency of such high-risk gene mutations was lower in this healthy donor cohort than in cross-sectional studies,^8,9^ which could either reflect their association with clinical abnormalities that would lead to exclusion from the donor pool or a difference in age structure of donors compared with population-based studies of CH.^24^

A recommendation to incorporate systematic screening for CH into the standard evaluation of transplant donor candidates would require synthesis of scientific evidence, technical feasibility, cost effectiveness, and ethical considerations.^18,19^ Our results provide scientific evidence for such a policy discussion by showing that individuals age 40 or older should not be excluded from stem cell donation based on identification of clonal hematopoiesis involving sole *DNMT3A* or *TET2* mutations. In contrast, clinicians may consider excluding individuals with splicing factor or *TP53* mutated CH, or with germline predisposition alleles, from stem cell donation due to an apparently elevated risk of donor cell leukemia. By exonerating donors with the most common form of clonal hematopoiesis, this study expands the older donor pool for patients unable to find HLA matched younger donors in unrelated registries or in whom matched sibling donors are preferred.

## Data Availability

The authors declare that all genetic data supporting the findings of this study are available within the paper and the supporting supplementary materials. Deidentified clinical and genomic data are available from the corresponding author upon reasonable request.

## Acknowledgements

This work was supported by the National Institutes of Health [K08CA204734 (RCL), P01CA229092 (RCL, JR, RJS), K08HL136894 (LPG), R01HL156144 (LPG)], the Damon Runyon Cancer Research Foundation (CJG), the Alan and Lisa Dynner Fund (RS, RCL), the James A. and Lois J. Champy Family Fund (RCL), the Jock and Bunny Adams Education and Research Fund (JHA), the Ted and Eileen Pasquarello Tissue Bank in Hematologic Malignancies, and the Connell and O’Reilly Families Cell Manipulation Core Facility.

## Notes

### Competing Interest Statement

The authors have declared no competing interest.

### Funding Statement

National Institutes of Health K08CA204734 (RCL), P01CA229092 (RCL, JR, RJS), K08HL136894 (LPG), R01HL156144 (LPG), Damon Runyon Cancer Research Foundation (CJG), the Alan and Lisa Dynner Fund (RS, RCL), the James A. and Lois J. Champy Family Fund (RCL), the Jock and Bunny Adams Education and Research Fund (JHA)

### Author Declarations

This study was conducted with approval of the Dana-Farber Cancer Institute IRB (protocol #16-582) and the Johns Hopkins University IRB (IRB00141250)

## References

1. Schlenk, R. F. et al. Mutations and treatment outcome in cytogenetically normal acute myeloid leukemia. N. Engl. J. Med. 358, 1909–1918 (200).

2. Nakamura, R. et al. A Multi-Center Biologic Assignment Trial Comparing Reduced Intensity Allogeneic Hematopoietic Cell Transplantation to Hypomethylating Therapy or Best Supportive Care in Patients Aged 50-75 with Advanced Myelodysplastic Syndrome: Blood and Marrow Transplant Clinical Trials Network Study 1102. Blood vol. 136 19–21 (2020).

3. Age requirements and limits for donating bone marrow. https://bethematch.org/transplant-basics/matching-patients-with-donors/why-donor-age-matters/.

4. Kollman, C. et al. The effect of donor characteristics on survival after unrelated donor transplantation for hematologic malignancy. Blood 127, 260–267 (2016).

5. Gragert, L. et al. HLA match likelihoods for hematopoietic stem-cell grafts in the U.S. registry. N. Engl. J. Med. 371, 339–348 (2014).

6. Pulsipher, M. A. et al. Related peripheral blood stem cell donors experience more severe symptoms and less complete recovery at one year compared to unrelated donors. Haematologica 104, 844–854 (2019).

7. McCurdy, S. R. et al. Effect of donor characteristics on haploidentical transplantation with posttransplantation cyclophosphamide. Blood Adv 2, 299–307 (2018).

8. Jaiswal, S. et al. Age-Related Clonal Hematopoiesis Associated with Adverse Outcomes. N. Engl. J. Med. (2014).

9. Genovese, G. et al. Clonal hematopoiesis and blood-cancer risk inferred from blood DNA sequence. N. Engl. J. Med. 371, 2477–2487 (2014).

10. Jaiswal, S. et al. Clonal Hematopoiesis and Risk of Atherosclerotic Cardiovascular Disease. N. Engl. J. Med. NEJMoa1701719 (2017).

11. Young, A. L., Challen, G. A., Birmann, B. M. & Druley, T. E. Clonal haematopoiesis harbouring AML-associated mutations is ubiquitous in healthy adults. Nat. Commun. 7, 12484 EP–.

12. Gibson, C. J. et al. Donor-engrafted CHIP is common among stem cell transplant recipients with unexplained cytopenias. Blood (2017) doi:10.1182/blood-2017-01-764951.

13. Frick, M. et al. Role of Donor Clonal Hematopoiesis in Allogeneic Hematopoietic Stem-Cell Transplantation. J. Clin. Oncol. 37, 375–385 (2019).

14. Wong, W. H. et al. Engraftment of rare, pathogenic donor hematopoietic mutations in unrelated hematopoietic stem cell transplantation. Sci. Transl. Med. 12, (2020).

15. Boettcher, S. et al. Clonal hematopoiesis in donors and long-term survivors of related allogeneic hematopoietic stem cell transplantation. Blood 135, 1548–1559 (2020).

16. Oran, B. et al. Donor clonal hematopoiesis increases risk of acute graft versus host disease after matched sibling transplantation. Leukemia (2021) doi:10.1038/s41375-021-01312-3.

17. Newell, L. F. et al. Engrafted Donor-Derived Clonal Hematopoiesis after Allogenic Hematopoietic Cell Transplantation is Associated with Chronic Graft-versus-Host Disease Requiring Immunosuppressive Therapy, but no Adverse Impact on Overall Survival or Relapse. Transplantation and Cellular Therapy vol. 27 662.e1–662.e9 (2021).

18. DeZern, A. E. & Gondek, L. P. Stem cell donors should be screened for CHIP. Blood Adv 4, 784–788 (2020).

19. Gibson, C. J. & Lindsley, R. C. Stem cell donors should not be screened for clonal hematopoiesis. Blood Adv 4, 789–792 (2020).

20. Seftel, M. D. et al. Clonal Hematopoiesis in Related Allogeneic Transplant Donors: Implications for Screening and Management. Biol. Blood Marrow Transplant. 26, e142– e144 (2020).

21. Sorror, M. L. et al. Hematopoietic cell transplantation (HCT)-specific comorbidity index: a new tool for risk assessment before allogeneic HCT. Blood 106, 2912–2919 (2005).

22. Armand, P. et al. A disease risk index for patients undergoing allogeneic stem cell transplantation. Blood 120, 905–913 (2012).

23. Abelson, S. et al. Prediction of acute myeloid leukaemia risk in healthy individuals. Nature 559, 400–404 (2018).

24. Malcovati, L. et al. Clinical significance of somatic mutation in unexplained blood cytopenia. Blood 129, 3371–3378 (06 22, 2017).

25. Eto, M. et al. Specific destruction of host-reactive mature T cells of donor origin prevents graft-versus-host disease in cyclophosphamide-induced tolerant mice. J. Immunol. 146, 1402–1409 (1991).

26. Wachsmuth, L. P. et al. Post-transplantation cyclophosphamide prevents graft-versus-host disease by inducing alloreactive T cell dysfunction and suppression. J. Clin. Invest. 129, 2357–2373 (2019).

27. Jeong, M. et al. Large conserved domains of low DNA methylation maintained by Dnmt3a. Nat. Genet. 46, 17–23 (2014).

28. Ley, T. J. et al. DNMT3A mutations in acute myeloid leukemia. N. Engl. J. Med. 363, 2424– 2433 (2010).

29. Walter, M. J. et al. Recurrent DNMT3A mutations in patients with myelodysplastic syndromes. Leukemia 25, 1153–1158 (2011).

30. Patel, J. P. et al. Prognostic relevance of integrated genetic profiling in acute myeloid leukemia. N. Engl. J. Med. 366, 1079–1089 (2012).

31. Russler-Germain, D. A. et al. The R882H DNMT3A mutation associated with AML dominantly inhibits wild-type DNMT3A by blocking its ability to form active tetramers. Cancer Cell 25, 442–454 (2014).

32. Miles, L. A. et al. Single-cell mutation analysis of clonal evolution in myeloid malignancies. Nature (2020) doi:10.1038/s41586-020-2864-x.

33. Young, A. L., Tong, R. S., Birmann, B. M. & Druley, T. E. Clonal hematopoiesis and risk of acute myeloid leukemia. Haematologica 104, 2410–2417 (2019).

34. Kim, S. J. et al. A DNMT3A mutation common in AML exhibits dominant-negative effects in murine ES cells. Blood 122, 4086–4089 (2013).

35. Buscarlet, M. et al. Lineage restriction analyses in CHIP indicate myeloid bias for TET2 and multipotent stem cell origin for DNMT3A. Blood blood–2018–01–829937 (2018).

36. Nisticò, A. & Young, N. S. gamma-Interferon gene expression in the bone marrow of patients with aplastic anemia. Ann. Intern. Med. 120, 463–469 (1994).

37. Baldridge, M. T., King, K. Y., Boles, N. C., Weksberg, D. C. & Goodell, M. A. Quiescent haematopoietic stem cells are activated by IFN-gamma in response to chronic infection. Nature 465, 793–797 (2010).

38. Young, K. et al. Decline in IGF1 in the bone marrow microenvironment initiates hematopoietic stem cell aging. Cell Stem Cell (2021) doi:10.1016/j.stem.2021.03.017.

39. Gondek, L. P. et al. Donor cell leukemia arising from clonal hematopoiesis after bone marrow transplantation. Leukemia 30, 1916–1920 (2016).

40. Kobayashi, S. et al. Donor cell leukemia arising from preleukemic clones with a novel germline DDX41 mutation after allogenic hematopoietic stem cell transplantation. Leukemia vol. 31 1020–1022 (2017).

41. Nevejan, L. et al. Malignant progression of donor-engrafted clonal hematopoiesis in sibling recipients after stem cell transplantation. Blood advances vol. 4 5631–5634 (2020).

42. Yoshida, K. et al. Frequent pathway mutations of splicing machinery in myelodysplasia. Nature 478, 64–69 (2011).

43. Polprasert, C. et al. Inherited and Somatic Defects in DDX41 in Myeloid Neoplasms. Cancer Cell 27, 658–670 (2015).

44. Macatonia, S. E. et al. Dendritic cells produce IL-12 and direct the development of Th1 cells from naive CD4+ T cells. J. Immunol. 154, 5071–5079 (1995).

45. Christopher, M. J. et al. Immune Escape of Relapsed AML Cells after Allogeneic Transplantation. N. Engl. J. Med. 379, 2330–2341 (2018).

46. Murphy, E. E. et al. B7 and interleukin 12 cooperate for proliferation and interferon gamma production by mouse T helper clones that are unresponsive to B7 costimulation. J. Exp. Med. 180, 223–231 (1994).

47. Reddy, V. et al. Interleukin 12 is associated with reduced relapse without increased incidence of graft-versus-host disease after allogeneic hematopoietic stem cell transplantation. Biol. Blood Marrow Transplant. 11, 1014–1021 (2005).

48. Darlak, K. A. et al. Enrichment of IL-12-producing plasmacytoid dendritic cells in donor bone marrow grafts enhances graft-versus-leukemia activity in allogeneic hematopoietic stem cell transplantation. Biol. Blood Marrow Transplant. 19, 1331–1339 (2013).

49. Kernan, N. A. et al. Clonable T lymphocytes in T cell-depleted bone marrow transplants correlate with development of graft-v-host disease. Blood 68, 770–773 (1986).

50. Ghoneim, H. E. et al. De Novo Epigenetic Programs Inhibit PD-1 Blockade-Mediated T Cell Rejuvenation. Cell 170, 142–157.e19 (2017).

51. Youngblood, B. et al. Effector CD8 T cells dedifferentiate into long-lived memory cells. Nature 552, 404–409 (2017).

52. Herold, S. et al. Donor cell leukemia: evidence for multiple preleukemic clones and parallel long term clonal evolution in donor and recipient. Leukemia 31, 1637–1640 (2017).

53. Rojek, K. et al. Identifying Inherited and Acquired Genetic Factors Involved in Poor Stem Cell Mobilization and Donor-Derived Malignancy. Biol. Blood Marrow Transplant. (2016) doi:10.1016/j.bbmt.2016.08.002.

